# Reactions to an Online Demonstration of the Effect of Increased Fruit and Vegetable Consumption on Appearance: Survey

**DOI:** 10.1101/19004523

**Authors:** Patrick Cairns, Gozde Ozakinci, David Ian Perrett

**Affiliations:** Perception Lab, School of Psychology & Neuroscience, University of St Andrews, St Mary’s Quad, South Street, St Andrews, Scotland, KY16 9JP, UK; School of Medicine, Medical & Biological Sciences Building, University of St Andrews, North Haugh, St Andrews, Scotland, KY16 9TF UK

**Keywords:** Diet, skin appearance, motivations, fruit and vegetables, carotenoid

## Abstract

**Background:** Inadequate fruit and vegetable consumption causes a considerable disease burden and premature mortality. Despite considerable public health promotion of a healthy diet the average consumption is still below recommended levels. Fruit and vegetable consumption influences human skin colour, increasing red/yellow/orange pigment in the skin. Given that this colour is deemed attractive and healthy-looking, the appearance benefit may provide motivation to eat more fruit and vegetables. Such appearance motivation could be particularly effective in young individuals who currently eat least fruit and vegetables.

**Objectives:** To assess how widely the impact of diet on skin colour is known within the UK. To compare the strength of motivation to eat fruit and vegetables based on health and appearance benefits and to compare the effect of different UK demographics on motivation.

**Methods:** Four groups of UK residents (N = 200 each group) were recruited through the Prolific online platform. Groups comprised younger (aged 18-24) and older adults (aged 40-60) of low and high self-reported socioeconomic status (1-5 and 6-10 on a 10-point rating scale). Facial images simulating the skin colour associated with low and high fruit and vegetable diets were shown to participants. Questionnaires were used to assess (1) background knowledge of the health and skin colour effects of dietary fruit and vegetables; (2) the specific motivational impact of the skin colour illustration and (3) the relative importance of motivation to consume fruit and vegetables arising from health and skin colour appearance benefits.

**Results:** (1) 61% of all participants were unaware of the dietary–skin colour association. (2) 57% of participants found the simple demonstration of the dietary impact on skin colour positively motivating to eat more fruit and vegetables. The visual demonstration was equally motivating for participants of high and low self-reported socioeconomic status (*P* = .63) and different ethnic backgrounds (White N = 453, Black N = 182, Asian N = 87, *P* = .22). Health benefits from a diet high in fruit and vegetables were regarded as more motivating than skin colour appearance benefits. The appearance benefits of a high fruit and vegetable diet (compared to the health benefits) were relatively more important for the younger participants (Mann-Whitney U = 96,263, *P* < .001) and for women (N = 489) than for men (N = 310, U = 83,763, *P* = .01).

**Conclusions:** These findings indicate that promotion of the skin colour effects of diets high in fruit and vegetables could provide additional motivation for a healthier diet. Our study indicates the wide appeal of appearance benefits from dietary fruit and vegetable (across ethnicity and socioeconomic status) and particularly amongst young adults where inadequate diet is most prevalent.

## Introduction

Inadequate fruit and vegetable consumption is estimated to lead to between 5.6 to 7.8 million premature deaths per year worldwide [1], chiefly through incidences of cardiovascular disease including coronary heart disease and stroke [1,2,3], diabetes and its complications [4,5] and several cancers [1,2,6]. Globally inadequate intake of fruit and vegetables is also responsible for the loss of up to 103 million years of healthy life due to disability [7]. The negative ramifications of these lifestyle-attributable diseases are widely felt. In addition to the consequences for personal wellbeing, poor population health contributes to an overburdening of healthcare systems and fiscal strain due to lost productivity [8]

Only 29% of adults in the UK [9] report eating the recommended 5 portions of fruit and vegetables per day. Fewer men (26%) than women (32%) meet the 5 a day guideline. Young people aged 16-24 were also less likely than other adults aged 45-60 to get their 5 a day (average 3.3 portions per day compared to 3.8). Figures are worse for children with only 18% of children aged 5-15 eating 5 portions per day. Residential area deprivation and lower socioeconomic status independently predict decreased fruit and vegetable consumption [10,11].

Health promotion efforts to increase fruit and vegetable consumption vary from individual approaches such as personal (or parental) advice and counseling to public health campaigns. Although much has been achieved in terms of understanding what psychological techniques may work or not to facilitate healthy eating [12], given the current state of inadequate consumption of fruit and vegetables, we are still in need of novel and innovative methods in addition to existing ones to encourage healthy eating.

### Skin colour effects

Fruit and vegetable consumption influences human skin colour, increasing the presence of red/yellow carotenoid pigments in the skin [13-17]. Indeed, this skin colour is a reliable biomarker of fruit and vegetable and carotenoid intake [18-21]. When asked to choose the skin colour that looks healthiest in photographs of themselves or others, participants consistently choose a skin colour which represents a higher fruit and vegetable consumption [13,22-24]. Likewise when asked to choose the most attractive facial photograph, participants choose the one with increased carotenoid skin coloration over baseline coloration [25] and even over increased suntan coloration [26,27]. Some reports point to a limitation of the effect of carotenoid skin colour.

Appleton et al. [28] found that carotenoid colour did not affect attractiveness when pose and expression were unconstrained, and Tan et al. [23] found that while a subtle level of carotenoid colour increased attractiveness a large amount was deemed unattractive in a Malaysian Chinese population.

In terms of using the effect of fruit and vegetable consumption on skin colour to motivate increased fruit and vegetable consumption, limited trials have been conducted. Whitehead et al. [29] found that those shown a personalised demonstration of the potential skin colour improvements arising from increased fruit and vegetable consumption reported an increase in fruit and vegetable consumption at a 10-week follow-up.

The present study aimed to assess the extent to which the effects of fruit and vegetable consumption on skin colour are known within the UK. After 7 years of published literature and extensive media coverage, we expected that the effect of fruit and vegetable consumption on skin colour will be known by a proportion of the population but the demographics of those familiar with the effect are unclear.

Secondly, we aimed to assess the extent to which UK residents are motivated to eat fruit and vegetables by a simple demonstration of the effects of fruit and vegetable consumption on skin colour. Given the previous studies, we expected that a majority of individuals would express positive motivation following the online demonstration.

Certain subsections of society may be more receptive to appearance-based incentives than others. Within the USA, Hayes and Ross [30] found younger participants to be more concerned about their appearance than older participants. Women were also more concerned about their appearance than men. Across all demographic groups appearance was a powerful motivation for healthy eating [30]. Chung et al. [31] found appearance to be an important factor in women’s decisions to eat fruit and vegetables. It was therefore predicted that younger participants and female participants would show the greatest motivation to increase fruit and vegetable consumption following the online demonstration. While it is clear that low socioeconomic status is predictive of reduced fruit and vegetable consumption, it is not known whether socioeconomic status relates to the dietary motivation from skin colour.

## Methods

Ethical approval was obtained from University of St Andrews School of Psychology Ethics Committee (PS13092). 802 participants completed testing, mean age = 35.2 (SD = 14.2). 489 females (61%) and 311 males (38.8%) participated. Participants were recruited via Prolific, an online recruitment platform in four waves: (1) 202 18-26 year-olds who rated themselves as belonging to the lower half of self-perceived socioeconomic status (rating of 1-5 on a 10 point visual analogue ladder scale), (2) 199 18-26 year-olds who rated themselves as belonging to the upper half of self-perceived socioeconomic status (6-10 rating on the 10 point scale), (3) 200 40-60 year-olds who rated themselves as belonging to the lower half of self-perceived socioeconomic status, and (4) 201 40-60 year-olds who rated themselves as belonging to the upper half of self-perceived socioeconomic status.

Participants were paid £0.6 for taking part (at an average rate of £9 per hour), with the questionnaire taking 4 minutes approximately to complete.

Image transforms. Participants were shown a before and after photo of a male face (Figure 1). This photo had been graphically changed to represent the skin colour changes that occur following the consumption of five portions of fruit and vegetables for 6 weeks. This single image pair was used to make the demonstration as simple as possible, and as the skin colour effect of fruit and vegetable consumption is the same for men and women. Images were transformed to simulate the effect of increasing dietary fruit and vegetables using methods outlined previously [13,22,23,26].

The colour change in the presented image pair was measured by selecting a large patch of the image of facial skin between the eyes and neck and analysing the average colour values of the pixels within the image patch. This showed that the right hand image in Figure 1 was darker, redder and yellower than the left image. The colour difference between right and left images in CIELab colour space was L*a*b*=-1.5, 2.0, 4.1. The displayed colour difference is equivalent to the skin colour change (L*a*b*=-1.8, 1.2, 3.8) after 4 weeks of a 500ml/day dietary smoothie supplement of 6 extra daily portions of fruit and vegetables [14]. The colour difference we use is one quarter of the difference used in other studies [26,32].

Figure 1. [Omitted from preprint, please email authors to access Figure 1]

### Procedure

Participants were first shown an image (Figure 1) showing the effect of fruit and vegetable consumption on skin colour benefits. Participants were then given a questionnaire asking demographics (gender, age, ethnicity) and how many portions of fruit and vegetables they eat per day, ranging from 0 to 10 or more. They were also asked whether they were aware that diet affected skin colour appearance and health. “Before seeing this display were you aware that eating fruit and vegetables (a) can impart a golden glow to your skin colour? (b) may help reduce the risk of the two main killer diseases in the UK - heart disease and some cancers?”. Answer options were: no (0), somewhat (1), yes (2).

One question assessed the motivational effect of the display: “Has seeing the effect of diet on skin colour in this exhibit made you want to eat less or more fruit and vegetables?”. Answer options were: a lot less, a little less, no change, a little more, a lot more.

Two further questions assessed the relative motivation of participants: “How much would the following make you want to eat more fruit and vegetables: (a) getting a skin colour that looks healthy and attractive (within 20 days), (b) reducing your chance of heart disease and some forms of cancer?” Answer options were: none at all, a little, a moderate amount, a lot, a great deal.

### Analysis

Non-parametric statistics were used for ordinal data (awareness and motivation) and for fruit and vegetable consumption as skew departed from normality.

## Results

### Demographics of participants

Gender. 489 females (61%) and 311 males (38.8%) took part. Two individuals reported their gender as ‘Other’.

Age. 381 younger participants (50.26%) reported age between 18-30 years (mean 21.57, SD +/- 2.09). 385 older participants (49.74%) reported age between 38-61 years (48.7 +/- 5.83).

#### Socioeconomic status

We sampled individuals with self-perceived socioeconomic status scale ranging from 1 (the lowest) to 10 (the highest), mean rating = 5.32 (SD = 1.67). The frequency of self-report was 12, 32, 72, 123, 157, 190, 148, 47, 4, and 5 across the 10 status levels.

#### Ethnicity

Participants were asked to choose an image which best represented their ethnic background from a choice of 4 images representing Black (African), White (Caucasian), East Asian and West Asian (Indian/Pakistani). 182 participants (22.7%) identified themselves as having African ethnicity, 87 participants (10.8%) as Asian, 454 participants (56.8%) as Caucasian, 23 participants (2.9%) as West Asian, 36 participants (4.5%) as having more than one ethnicity, and 18 participants (2.2%) an ethnicity not shown by our questionnaire stimuli.

#### Fruit and vegetable consumption

The mean level of fruit and vegetable consumption across the entire sample was 3.38 +/- 1.76 (mean +/- SD, skew 0.40 and kurtosis = 0.10) portions per day. Young adults reported eating less than older adults (3.13 +/- 1.72 versus 3.63 +/- 1.76 portions per day; younger adults mean rank = 363.9, N = 399; older adults mean rank = 436.9, N = 401; Mann-Whitney U = 94,601, *P* < .001, r = .161). Low socioeconomic status participants reported eating less than those of high socioeconomic status (3.15 +/- 1.70 versus 3.62 +/- 1.78 portions per day respectively; low status mean rank = 363.45, N = 395; high status mean rank = 425.71, N = 393; U = 89,884, *P* < .001, r = .139). Men reported eating less than women (3.11 +/- 1.60 versus 3.55 +/- 1.83 portions per day; men mean rank = 367.25, N = 310; women mean rank = 419.99, N = 488; U = 85,638, *P* < .001, r = .114). Consumption of fruit and vegetables did not differ across three ethnic groups, Black, White and Asian (N = 721, Kruskal-Wallis Test = 1.884, df = 2, *P* = .39).

### Awareness of appearance benefits

487 participants (61%) were unaware of the association between diet and skin colour; 219 (27%) were somewhat aware, and 95 (12%) were aware. This contrasts with the 66 (8.2%) participants who were unaware of health risks (e.g. cancer, heart disease) of a diet low in fruit and vegetables. Younger participants were more aware of the fruit and vegetable effect on skin colour than older participants (younger adults mean rank = 417.67, N = 400; older adults mean rank = 384.37, N = 401; U = 73,531, *P* = .019, r = - .081). Participants reporting low socioeconomic status were equally aware of the effect than participants reporting high socioeconomic status (low socioeconomic status mean rank = 397.13, N = 396; high socioeconomic status mean rank = 392.86, N = 393; U = 76,972, *P* = .762, r = -.01). Women were more aware of the fruit and vegetable effect on skin colour than men (women mean rank = 416.67, N = 489, men mean rank = 373.71, N = 310; U = 83,946, *P* = .003, r = .104). Awareness of the fruit and vegetable effect on skin colour differed across three ethnic groups, Black, White and Asian (N = 722, Kruskal-Wallis Test = 7.915, df = 2, *P* = .019). White participants showed least awareness (Black mean rank = 382.71, Asian mean rank = 392.55, White mean rank = 347.02),

### Motivation change from seeing effect on skin colour

457 participants (57%) reported the simple demonstration of the dietary impact on skin colour positively motivating to eat more fruit and vegetables.

#### Age

Comparing younger (18-26 years old) and older participants (40-60 years old) showed that the younger group was more likely to endorse the view that the appearance demonstration motivated diet change than the older group (Figure 2, younger participants mean rank = 435.79, N = 400; older participants mean rank = 366.30, N = 401; U = 66,284, *P* < .001, r = -.164).

**Figure 2.**
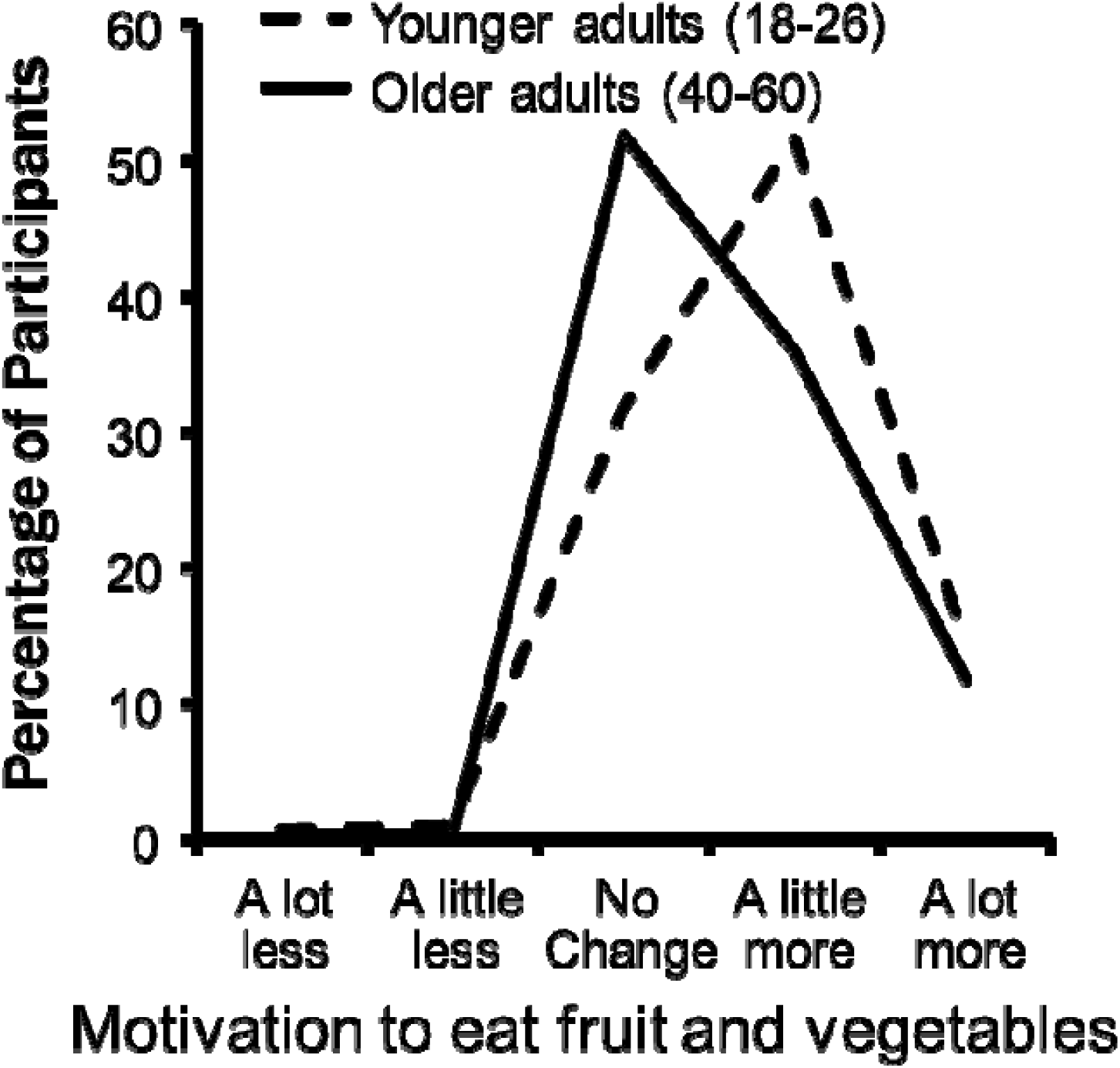
Effect of age on appearance motivation. Change in motivation to eat fruit and vegetables as a result of seeing the demonstration images for two age groups. Younger adults were more positively motivated compared to older participants.

#### Gender

Participant gender had no significant effect on the motivation for diet change from the demonstration of the effects of fruit and vegetables on skin colour (women mean rank = 408.37, N = 489, men mean rank = 386.80, N = 310; U = 79,886, *P* = .16, r = .05). The appearance demonstration was thus equally motivating to women and men. Socioeconomic status. Comparing low (self-reporting levels 1-5) and high socioeconomic status (self-reporting levels 6-10) participants showed that the two groups were equally likely to endorse the view that the appearance demonstration was motivating to change diet (mean rank low = 396.12, N = 396; mean rank high = 393.88, N = 393; U = 77,372, *P* = .880, r = -.005). Hence, the appearance motivation was equal across socioeconomic status.

#### Ethnicity

The distribution of answers (to the question of whether the demonstration of skin colour change motivated dietary change) across three ethnic groups, Black, White and Asian, was not significantly different (N = 721, Kruskal-Wallis Test = 2.999, df = 2, *P* = .22). Comparing the median score of participants answering this question to a median of zero, the score expected by random choice, each ethnic group showed a significant bias to answer the question affirmatively (Black participants: One Sample Wilcoxon Signed Rank test T = 5,899.5, N = 182, asymptotic two-tailed significance *P* < .001; Asian participants: T = 1,573.5, N = 87, *P* < .001; White participants: T = 31,311, n = 452, *P* < .001). Hence Black, White and Asian groups found the colour demonstration motivating (Figure 1).

### Appearance vs Health Motivation for dietary fruit and vegetables

Ninety percent of participants answered that skin colour appearance would provide motivation to eat more fruit and vegetables. By contrast virtually all participants (99%) were motivated to a greater or lesser extent by the health benefits of fruit and vegetables in reducing the chance of heart disease and cancer (Figure 3). To compare the relative importance of health and appearance motivation the response categories were recoded as a score (0-4). The mean numerical score for health motivation was 3.27 while the mean score for appearance was 2.30. Wilcoxon Signed Rank test T = 118,103, N = 801, *P* < .001, r = .618).

**Figure 3.**
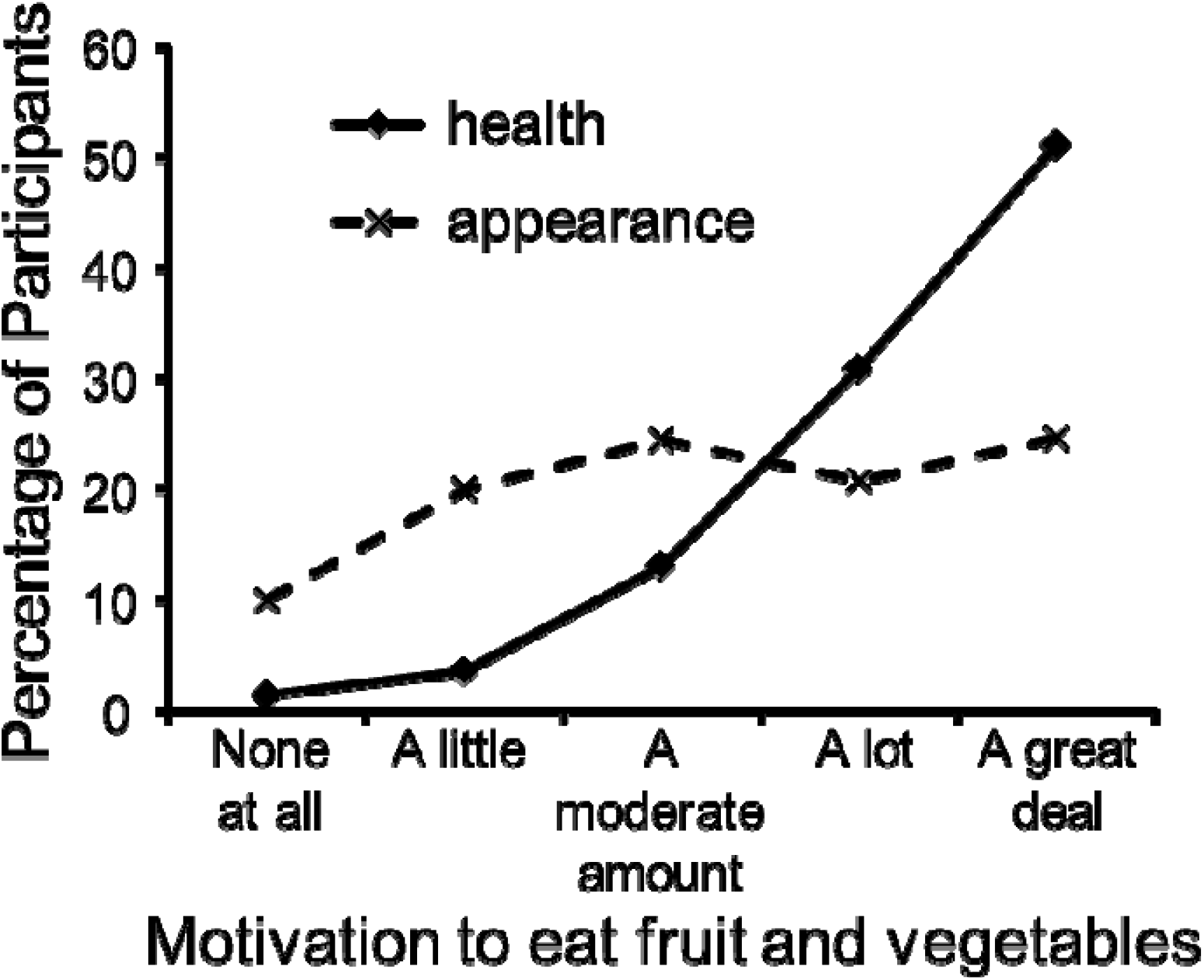
Relative motivation of health and appearance for consumption of fruit and vegetables.

#### Gender

To compare health and appearance motivation an appearance/health contrast score was computed for each participant (the 0 - 4 score of the importance of appearance minus the 0 - 4 score of the importance of health for consuming fruit and vegetables). Comparing contrast scores for women and men showed that the relative importance of appearance to health in motivating diet change was greater for women (mean rank = 416.29, N = 489) than men (mean rank = 374.3, N = 310, U = 83,763, *P* = .01, r = .09).

#### Age

Younger adults (mean rank = 442.96, N = 400) found the appearance benefits more motivating relative to the health benefits (mean rank = 359.15, N = 401, U = 63,416, *P* < .001, r = .19).

#### Socioeconomic status

There was no effect of socioeconomic status on the relative motivation of health and appearance (mean rank high = 392.52 N=393 mean rank low = 397.46 N = 396, U = 78,839, *P* = .751, r = -.011).

#### Knowledge of the fruit and vegetables on skin colour

The importance of appearance relative to health in motivating diet change was greater for those participants who already knew that diet alters skin colour (mean rank = 445.79, N = 314) than for those not knowing the effect (mean rank = 372.12, N = 487, U = 90,522, *P* < .001, r = .161).

## Discussion

The two major findings of the study were (a) the limited percentage of people (40%) with knowledge of the dietary–skin colour association and (b) that many people (57%) found the simple demonstration of the dietary impact on skin colour positively motivating to eat more fruit and vegetables, while very few (1.2%) reported the demonstration de-motivating. The combination of these two findings point to the benefits that can be accrued through publicising and demonstrating the dietary effect on skin colour. Media coverage in the UK has been extensive (reaching millions of TV viewers) but there is still a substantial proportion of the population that reports not knowing the dietary effects. Of the 801 participants, 487 (61%) were unaware of the effects of fruit and vegetable consumption on skin colour. Of these 248 (51%) stated they were motivated to eat more fruit and vegetables as a result of seeing the demonstration. Hence increasing public awareness of the dietary effects on skin could provide additional motivation for a healthy diet for 31% of the sample population (% of those unaware multiplied by % of those motivated). Extrapolating this to the UK adult population of approximately 50 million, this amounts to 16.5 million people.

### Age and socioeconomic status

It is noteworthy that the motivational effects of appearance were most prominent in the younger adults. It is this young adult section of society (particularly in the low socioeconomic bracket) that have the lowest fruit and vegetable consumption [9]. In the current study it was again evident that the younger adults and those reporting low socioeconomic status were reporting lower fruit and vegetable consumption. We were unable to detect any influence of socioeconomic status on appearance motivation. Hence, promotion of appearance benefit would not widen health inequalities. Indeed, drawing attention to the colour benefits of a healthy diet would have particular impact on a younger population, thereby increasing the chance for life-long adoption of an improved diet.

### Gender

We expected women to be more motivated by appearance benefits than men. Comparing reactions to the visual demonstration we found no significant impact of gender. For both men and women in the present study, motivation for a diet high in fruit and vegetables was greater for the benefits to health than the motivation for an improved skin colour appearance. Nonetheless motivation from benefits to appearance relative to benefits to health was higher in women than in men.

### Fruit and vegetable consumption

The average consumption of fruit and vegetables reported by the entire sample here was 3 portions per day (significantly less than the 5 a day recommended by the British National Healthcare System). Hence publicity of the appearance benefits of a good diet rich in fruit and vegetables could augment motivation for a healthy diet. It is relevant here that the majority of our participants (57%) reported that the appearance benefits were positively motivating.

### Ethnicity effects

The participants in this study reported a variety of ethnic backgrounds (23% Black, 11% Asian and 57% White). The effect of carotenoids is likely to be less apparent for individuals with darkly pigmented skin. Nonetheless, carotenoid effects on skin colour are apparent across different ethnicities. Coetzee & Perrett [33] found that carotenoid supplementation produced an increase in skin yellowness in sun-protected skin regions of African participants. Likewise a diet rich in fruit and vegetables increases skin yellowness in Asian participants [14,23] as it does in White participants [13,15,25]. While the skin colour effects of diet are detectable in a range of ethnic groups, different cultures may vary in their perception of whether the skin colour change is desirable. Black South African participants were found to perceive increased skin yellowness positively [22]. Yet a recent report indicates that Asian participants from Malaysia find only a slight increase in carotenoid skin colour more attractive than the skin colour baseline [23]. A further report concludes that mainland Chinese participants do not find increased yellow skin pigmentation attractive [34]. Nonetheless, whatever the influence of culture on colour preferences, the current study found that Asian, Black and White participants from the UK were equally impressed by the impact of diet on skin colour.

### Limitations

We have detected several factors associated with attitudes to the appearance benefit arising from a diet high in fruit and vegetables. Socioeconomic status was not found to have an effect on appearance motivation, but we may have confounded area of social deprivation with low socioeconomic status, since both of these factors can have an impact on dietary behaviour.

We illustrated the effect with a single Caucasian male face. The demonstration may be more effective if displayed with the same gender, age and ethnicity as the participant or better still with an image of the participant [29].

We measured the effects of appearance on self-reported motivation to consume more fruit and vegetables. We acknowledge that motivation is necessary but not sufficient for actual dietary change [35]. The impact of appearance on real dietary change is likely to be smaller than on motivation. Nonetheless any change in motivation to consume more fruit and vegetables is an important step in the right direction of a healthy lifestyle.

## Data Availability

Data uploaded as supplementary file.

## Acknowledgements

This work was supported by grants from a British Academy Wolfson Research Professorship (grant number: WRP/2008/87) and University of St Andrews Knowledge Exchange and Impact Fund 2017/18. The authors gratefully acknowledge Lesley Ferrier, Xue Lei and Martha Lucia Borras Guevara for assistance with study preparation, and Anne Perrett for proofreading.

## Financial Disclosure

None.

## Non-financial Disclosure

None.

## Notes

### Competing Interest Statement

The authors have declared no competing interest.

### Author Declarations

All relevant ethical guidelines have been followed and any necessary IRB and/or ethics committee approvals have been obtained.

Any clinical trials involved have been registered with an ICMJE-approved registry such as ClinicalTrials.gov and the trial ID is included in the manuscript.

